# Rare *PSAP* variants and possible interaction with *GBA* in REM sleep behavior disorder

**DOI:** 10.1101/2021.07.13.21258405

**Authors:** Yuri L. Sosero, Eric Yu, Mehrdad A. Estiar, Lynne Krohn, Kheireddin Mufti, Uladzislau Rudakou, Jennifer A. Ruskey, Farnaz Asayesh, Sandra B. Laurent, Dan Spiegelman, Jean-François Trempe, Timothy G. Quinnell, Nicholas Oscroft, Isabelle Arnulf, Jacques Y. Montplaisir, Jean-François Gagnon, Alex Desautels, Yves Dauvilliers, Gian Luigi Gigli, Mariarosaria Valente, Francesco Janes, Andrea Bernardini, Karel Sonka, David Kemlink, Wolfgang Oertel, Annette Janzen, Giuseppe Plazzi, Elena Antelmi, Francesco Biscarini, Michela Figorilli, Monica Puligheddu, Brit Mollenhauer, Claudia Trenkwalder, Friederike Sixel-Döring, Valérie Cochen De Cock, Christelle Charley Monaca, Anna Heidbreder, Luigi Ferini-Strambi, Femke Dijkstra, Mineke Viaene, Beatriz Abril, Bradley F. Boeve, Ronald B. Postuma, Guy A. Rouleau, Abubaker Ibrahim, Ambra Stefani, Birgit Högl, Michele T.M. Hu, Ziv Gan-Or

**Affiliations:** Department of Human Genetics, McGill University, Montréal, QC, Canada; Montreal Neurological Institute, McGill University, Montréal, QC, Canada; Department of Neurology and Neurosurgery, McGill University, Montréal, QC, Canada; Department of Pharmacology & Therapeutics and Centre de Recherche en Biologie Structurale, McGill University, Montréal, Québec, Canada; Royal Papworth Hospital NHS Trust, Cambridge, UK; Sleep Disorders Unit, Sorbonne University, Institut du Cerveau - Paris Brain Institute - ICM, Inserm, CNRS, AP-HP, Hôpital de la Pitié Salpêtrière, Paris, France; Centre d’Études Avancées en Médecine du Sommeil, Hôpital du Sacré-Cœur de Montréal, Montréal, QC, Canada; Department of Psychiatry, Université de Montréal, Montréal, QC, Canada; Department of Psychology, Université du Québec à Montréal, Montréal, QC, Canada; Department of Neurosciences, Université de Montréal, Montréal, QC, H3T 1J4, Canada; National Reference Centre for Orphan Diseases, Narcolepsy- Rare hypersomnias, Sleep Unit, Department of Neurology, CHU Montpellier, Institute for Neurosciences of Montpellier INM, Univ Montpellier, INSERM, Montpellier, France; Clinical Neurology Unit, Department of Neurosciences, University Hospital of Udine, Udine, Italy; Department of Medicine (DAME), University of Udine, Udine, Italy; Department of Neurology and Centre of Clinical Neuroscience, Charles University, First Faculty of Medicine and General University Hospital, Prague, Czech Republic; Department of Neurology, Philipps University, Marburg, Germany; Department of Biomedical, Metabolic and Neural Sciences, University of Modena and Reggio Emilia, Modena, Italy; IRCCS, Institute of Neurological Sciences of Bologna, Bologna, Italy; Neurology Unit, Movement Disorders Division, Department of Neurosciences, Biomedicine and Movement Sciences, University of Verona, Verona, Italy; Department of Biomedical and Neuromotor Sciences (DIBINEM), Alma Mater Studiorum, University of Bologna, Bologna, Italy; Department of Medical Sciences and Public Health, Sleep Disorder Research Center, University of Cagliari, Cagliari, Italy; Paracelsus-Elena-Klinik, Kassel, Germany; Department of Neurology, University Medical Centre Göttingen, Göttingen, Germany; Sleep and Neurology Unit, Beau Soleil Clinic, Montpellier, France; EuroMov, University of Montpellier, Montpellier, France; University Lille North of France, Department of Clinical Neurophysiology and Sleep Center, CHU Lille, Lille, France; Department of Sleep Medicine and Neuromuscular Disorders, University of Münster, Münster, Germany; Department of Neurological Sciences, Università Vita-Salute San Raffaele, Milan, Italy; Laboratory for Sleep Disorders, St. Dimpna Regional Hospital, Geel, Belgium; Department of Neurology, St. Dimpna Regional Hospital, Geel, Belgium; Department of Neurology, University Hospital Antwerp, Edegem, Antwerp, Belgium; Sleep disorder Unit, Carémeau Hospital, University Hospital of Nîmes, France; Department of Neurology, Mayo Clinic, Rochester, MN, USA; Sleep Disorders Clinic, Department of Neurology, Medical University of Innsbruck, Innsbruck, Austria; Oxford Parkinson’s Disease Centre (OPDC), University of Oxford, Oxford, United Kingdom; Nuffield Department of Clinical Neurosciences, University of Oxford, Oxford, United Kingdom

**Keywords:** REM sleep behavior disorder, *PSAP*, saposin C, Parkinson’s disease, *GBA*, glucocerebrosidase, genetics

## Abstract

*PSAP* encodes saposin C, the co-activator of glucocerebrosidase, encoded by *GBA*. Since *GBA* mutations are associated with idiopathic/isolated REM sleep behavior disorder (iRBD), a prodromal stage of synucleinopathy, we examined the role of *PSAP* mutations in iRBD. We fully sequenced *PSAP* and performed Optimized Sequence Kernel Association Test in 1,113 iRBD patients and 2,324 controls. We identified loss-of-function (LoF) mutations, which are very rare in *PSAP*, in three iRBD patients and none in controls (uncorrected *p*=0.018). Two variants were stop mutations, p.Gln260Ter p.Glu166Ter, and one was an in-frame deletion, p.332_333del. All three mutations have a deleterious effect on saposin C, based on *in silico* analysis. In addition, the two carriers of p.Glu166Ter and p.332_333del mutations also carried a *GBA* variant, p.Arg349Ter and p.Glu326Lys, respectively. The co-occurrence of these extremely rare *PSAP* LoF mutations in two (0.2%) *GBA* variant carriers in the iRBD cohort, is unlikely to occur by chance (estimated co-occurrence in the general population based on gnomAD data is 0.00035%). Although none of the three iRBD patients with *PSAP* LoF mutations have phenoconverted to an overt synucleinopathy at their last follow-up, all manifested initial signs suggestive of motor dysfunction, two were diagnosed with mild cognitive impairment and all showed prodromal clinical markers other than RBD. Their probability of prodromal PD, according to the Movement Disorder Society research criteria was 98% or more. These results suggest a possible role of *PSAP* variants in iRBD and potential genetic interaction with *GBA*, which requires additional studies.

## Introduction

Rapid eye movement (REM) sleep behavior disorder (RBD) is characterized by the enactment of dreams during the REM phase of sleep [1]. In its idiopathic/isolated form (iRBD, presenting before the clinical diagnosis of a neurodegenerative disease), it represents a common prodromal stage of synucleinopathies, including Parkinson’s disease (PD), dementia with Lewy bodies (DLB) and multiple system atrophy (MSA) [1, 2]. Notably, over 80% of iRBD cases convert to a synucleinopathy within 10-15 years [2, 3]. In line with their clinical overlap, iRBD and overt synucleinopathies also share some of their genetic risk factors. For example, iRBD and PD are both associated with *GBA* variants, which represent one of the most common genetic risk factors for both diseases [4, 5]. *GBA* variants display an incomplete penetrance in iRBD as well as in PD [4, 5], suggesting that other factors, genetic and/or environmental, contribute to the development of these disorders among *GBA* carriers.

*GBA* encodes glucocerebrosidase (GCase), a lysosomal hydrolase whose main function is the degradation of glucocerebrosides into ceramide and glucose, although it has additional substrates [5]. To properly function, GCase requires a co-activator, saposin C (sapC) [6]. This protein is one of the four active domains of a protein precursor, prosaposin, encoded by the *PSAP* gene. After its synthesis, prosaposin is cleaved by cathepsin D (CTSD) into its functional proteins: saposins A, B, C and D [7, 8]. Saposins are lysosomal cofactors that activate enzymes degrading sphingolipids. Mutations in *PSAP* have been associated with the accumulation of sphingolipids and with different lysosomal storage disorders (LSD). For example, Gaucher’s disease, an LSD that is typically caused by biallelic mutations in *GBA*, is also rarely caused by biallelic mutations in the sapC domain of *PSAP* [7, 9].

Whereas the association of *GBA* variants with PD is widely accepted, the role played by *PSAP* in general and sapC specifically in PD remains controversial. Studies in Asian populations suggested an association between *PSAP* variants and PD [10-14], yet these results did not replicate in Europeans [15-17]. These conflicting results may suggest a possible role played by ethnic differences and/or by the extreme rarity of deleterious *PSAP* variants, reducing their detection in PD. Despite the clinical, biological, and, possibly, genetic links of *PSAP* with *GBA* and PD, the role of *PSAP* in iRBD has not been investigated. Herein, we analyzed a multi-center cohort of 1,113 iRBD patients and 2,324 healthy controls to evaluate a possible association between rare *PSAP* variants and iRBD.

## Methods

### Population

The current study included 1,113 unrelated iRBD patients and 2,324 unrelated healthy controls of European descent. Details on the cohorts and their recruitment have been previously published [4]. RBD was diagnosed with video polysomnography (vPSG) according to the International Classification of Sleep Disorders, version 2/3 criteria [18, 19]. About 81% of the iRBD patients were males (N=897) and their mean age was 68 ± 9.4. Among the controls, 48% of the participants were males (N=1,122) and their mean age was 48 ± 16.7.

### Standard Protocol Approvals, Registrations, and Patient Consents

All patients signed an informed consent form before entering the study, and the study protocol was approved by the institutional review boards.

### Genetic analysis

The *PSAP* coding regions were fully sequenced using Molecular Inversion Probes (MIPs) as previously described [15, 20]. A detailed description of the MIPs library and protocols is available online (https://github.com/gan-orlab/MIP_protocol). Variant annotation was performed with ANNOVAR [21]. The frequency of each variant was extracted from the Genome Aggregation Database (gnomAD) [22]. Post-alignment quality control and variant calling were done using the Genome Analysis Toolkit (GATK, v3.8) [23] as previously described [24]. Full code is available at https://github.com/gan-orlab/MIPVar/.

### In silico structural analysis

The impact of the rare variants on the structure and function of the saposin chains was investigated with *in silico* structural analyses. The atomic coordinates of the human saposin chains B and C were downloaded from the Protein Data Bank [25](ID 1n69 and 1m12, respectively). Images were generated using PyMol v. 2.4.0.

### Statistical analysis

Rare *PSAP* variants were filtered using a minor allele frequency (MAF) threshold of < 0.01. To test for rare *PSAP* variants enrichment in iRBD patients we performed optimized sequence Kernel association test (SKAT-O) for all rare variants and subsets of rare variants. These subsets included nonsynonymous, regulatory, potentially functional (nonsynonymous, frameshift, stop-gain and splicing) and loss-of-function (frameshift, stop-gain and splicing) rare variants. A further subset consisted of variants predicted to have a high deleteriousness probability based on a Combined Annotation Dependent Depletion (CADD) score ≥ 12.37. SKAT-O analysis was performed using SKAT package in R 3.5.2 [26]. False discovery rate (FDR) correction was applied to correct for multiple comparisons, using Benjamini-Hochberg method with stats package in R 4.0.2.

## Results

We identified 59 rare variants within the *PSAP* region, of which 15 were nonsynonymous and 3 were loss of function (LoF) variants (Supplementary Table 1). The mean coverage was 568X, and a minimum threshold of 30X was applied for variant quality control. To evaluate if rare *PSAP* variants are associated with iRBD, we performed SKAT-O comparing iRBD patients and healthy controls. There was a nominally significant enrichment of rare *PSAP* LoF variants (*p*=0.018) in iRBD patients. However, after FDR correction, the results lost statistical significance (*p*=0.1, Table 1). Three out of 1,113 iRBD patients (0.3%) carried a rare *PSAP* LoF variant, while no carriers of LoF variants were found among the controls (0/2324, Supplementary Table 1). In particular, p.Gln260Ter and p.Glu166Ter are both stop variants located, respectively, within the sapB and between sapA and sapB domains, therefore the sapC domain is not translated. The p.332_333del mutation is an in-frame deletion located within the sapC domain.

**Table 1.** Optimized sequence Kernel association test (SKAT-O) for *PSAP* rare variants.

| **Rare variant subset** | **P.value** | **P.adj** |
| --- | --- | --- |
| **CADD** | 0.034792187 | 0.1043766 |
| **Encode** | 0.286264029 | 0.3021955 |
| **Func** | 0.302195501 | 0.3021955 |
| **LoF** | 0.017929809 | 0.1043766 |
| **NS** | 0.052448772 | 0.1048975 |
| **All** | 0.246565484 | 0.3021955 |

CADD: Variants selected based on a Combined Annotation Dependent Depletion threshold >12.37; Encode: variants in regulatory elements; Func: potentially functional variants; LoF; loss of function variants; NS: nonsynonymous; All: all rare variants; P.adj: corrected p-value for multiple comparisons using false discovery rate

Given the interplay between sapC and GCase, we examined whether any of these three iRBD patients with PSAP LoF mutations also carry a GBA variant. Furthermore, we tested the presence of GBA copy number variants (CNVs), as was done previously [27]. We found that two of the patients, carrying the p.Glu166Ter and p.332_333del variants, also carried a GBA variant: p.Arg349Ter and p.Glu326Lys, respectively. None carried GBA CNVs. All PSAP and GBA variants were confirmed by Sanger sequencing.

We further examined the frequency of *PSAP* LoF variants on gnomAD database v2.11 (https://gnomad.broadinstitute.org). None of the LoF variants found in this study have been reported in gnomAD, and the overall frequency of *PSAP* LoF variants was extremely low, with a total allele count of high-quality LoF variants of 10 in 141,456 individuals (∼0.007%, compared to ∼0.3% in the iRBD cohort). With a frequency of ∼5% in the general European population for *GBA* variants (based on gnomAD data), the estimated combined carrier frequency of both LoF *PSAP* variants and *GBA* variants is 0.00035%, compared to 0.2% observed in the iRBD cohort, more than a 500-fold difference.

### In silico structural analyses

To evaluate the impact of the three iRBD-associated variants on the structure and function of the saposin chains we performed *in silico* analyses. The p.Glu166Ter variant, located between saposin chains A and B, would result in the termination of expression for chains B-D. The p.Gln260Ter variant is located towards the C-terminus of the sapB domain and would result in the deletion of its C-terminal helix (Figure 1A), as well as in the termination of sapC and sapD translation. This deletion would also unfold sapB and prevent its dimerization, which is critical for binding lipids [28]. Finally, the variant p.332_333del is located in a linker between helices 1 and 2 of sapC (Figure 1B) [29]. This shortened linker would prevent the formation of stabilizing contacts between these helices and thus interfere with its ability to bind membranes and GCase. Therefore, all three variants result in a loss of function of the sapC chain.

**Figure 1.**
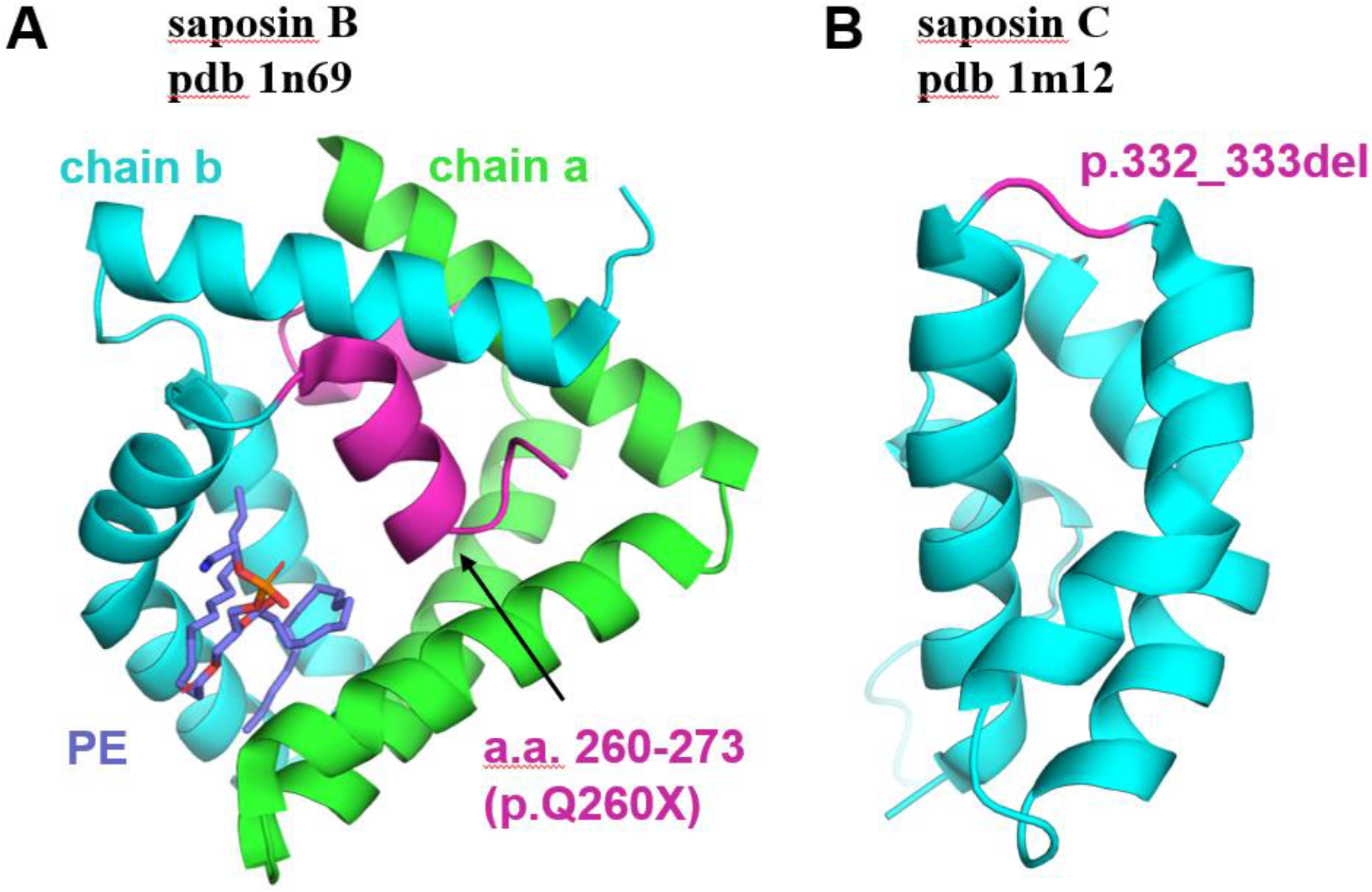
Structural analysis of the saposin B and C domains. (**A**) Crystal structure of the saposin B dimer (green and cyan, pdb 1n69). A bound phosphatidylethanolamine (PE) is shown in violet. The C-terminal helix (a.a. 260-273) deleted in the p.Gln260Ter variant is shown in magenta. (**B**) Solution NMR structure of the human saposin-C domain (cyan, pdb 1m12). In-frame deletion of amino acids Asn332 and Lys333 is shown in magenta.

### Clinical presentation of the iRBD patients with PSAP LoF variants

The iRBD patient with the p.332_333del *PSAP* variant was a male in the age range 75-79 who showed minor gait impairment, not quite erect posture, slight global slowness and poverty of spontaneous movements on the neurological examination. His Unified Parkinson’s Disease Rating Scale (UPDRS) III [30] score at last follow-up was 3. No cognitive deficits were present (Montreal Cognitive Assessment (MoCA) = 29/30), yet the patient manifested autonomic symptoms associated with prodromal PD, including constipation, erectile dysfunction and orthostatic hypotension. The risk of prodromal PD according to the Movement Disorder Society (MDS) research criteria [31] at the last follow-up was 1.000 (LR = 37452.7, Table 2).

**Table 2.** Clinical data at last follow-up of iRBD patients carrying *PSAP* rare variants.

| ***PSAP* LoF rare variants** | **p.332_333del** | **p.Glu166Ter** | **p.Gln260Ter** |
| --- | --- | --- | --- |
| ***GBA* variants** | p.Glu326Lys | p.Arg349Ter | No |
| **Sex** | Male | Male | Male |
| **AAD range** | 75-79 | 80-84 | 60-64 |
| **Disease duration** | >13 years | >2 years | >5 years |
| **Tremor** | No | No | No |
| **Hypokinesia** | No | Initial signs | No |
| **Bradykinesia** | No | Initial signs | Initial signs |
| **Postural instability** | No | No | No |
| **Cognitive symptoms** | No | Yes | Yes |
| **Psychiatric symptoms** | No | No | No |
| **Hyposmia** | No | Yes | Yes |
| **Orthostatic hypotension** | Yes | Yes | Dizziness standing up (negative tilt test) |
| **Constipation** | Yes | No | No |
| **Urinary dysfunction** | No | No | No |
| **Erectile dysfunction** | Yes | No | No |
| **Imaging signs** | Substantia nigra hyperechogenicity on the right side | / | / |
| **Risk prodromal PD** | 0.98 (288) – 1 (37458) | 0.96 (551) - 0.99 (25600) | 0.53 (88) - 0.98 (4072) |
| **MDS-UPDRS III** | 3 | 4 | 3 |
| **Smoker** | Yes (ex-smoker) | Yes (ex-smoker) | Yes (ex-smoker) |

LoF: loss of function variant; AAD range: age at diagnosis range; Disease duration: disease duration from age at diagnosis to last follow-up; Risk prodromal PD: risk for prodromal Parkinson's disease according to the Movement Disorder Society (MDS) criteria, not considering iRBD (values on the left) and considering iRBD (values on the right). Values in parentheses indicate the likelihood ratios; MDS-UPDRS: Movement Disorder Society - Unified Parkinson's Disease Rating Scale

The iRBD patient with the *PSAP* p.Glu166Ter variant was a male in the age range 80-84 displaying initial PD motor symptoms, including mild right leg rigidity, slight bilateral slowing of finger tapping movements and stooped posture, with a UPDRS III score of 4. He was also diagnosed with mild cognitive impairment (MCI, MoCA = 23/30). Furthermore, the patient had some non-motor PD-related symptoms, including significant hyposmia and orthostatic hypotension. His risk for prodromal PD was 0.99 (LR=25600, Table 2).

Finally, the iRBD patient with the *PSAP* p.Gln260Ter variant was a male in the age range 60-64 showing some signs of motor impairment, including mild asymmetric finger tapping and top tapping bradykinesia. His UPDRS III score was 3 and he was diagnosed with MCI (MoCA = 26/30). He displayed severe hyposmia, while no autonomic symptoms were present. His risk of prodromal PD at his last follow-up was 0.98 (LR=4072, Table 2).

## Discussion

In this study, we found three iRBD patients with extremely rare *PSAP* LoF variants, not reported on gnomAD, while no controls were found with LoF variants. Interestingly, two of the three *PSAP* LoF variant carriers also carried a *GBA* variant. While the enrichment of rare *PSAP* LoF variants in iRBD was only nominally significant, given their rarity it is plausible that this reflects a real association. Furthermore, assuming that in the general European population the carrier frequency of *GBA* variants is about 5%, and the carrier frequency of LoF variants (based on gnomAD) is about 0.007%, the probability to carry both a *GBA* variant and a *PSAP* LoF variant is 0.00035%. In the iRBD cohort, the carrier frequency of both was ∼0.2%, suggesting that this is likely not due to chance alone. The deleteriousness of the three *PSAP* LoF variants was further exemplified by structural analyses (Figure 1A and 1B). All iRBD patients met the MDS criteria for probable prodromal PD (Table 2).

Although the role of *PSAP* in iRBD and in synucleinopathies in general is still controversial, this study provides the first evidence for a possible role of *PSAP* variants in iRBD. The lack of a statistically significant enrichment in iRBD patients after correction for multiple comparisons can be explained by the extreme rarity of *PSAP* variants, resulting in insufficient power. The Residual Variation Intolerance Score (RVIS) of *PSAP* is -1, putting it in the top 8.47% of genes in the human genome which are intolerant to genetic variance, especially for LoF variance (FDR corrected *p*=0.00037 for the observed vs. expected number of LoF variants - http://genic-intolerance.org/Search?query=psap).

Two iRBD carriers of *PSAP* LoF variants were also carriers of a *GBA* variant. Given the incomplete penetrance of *GBA* in iRBD, the presence of potentially pathogenic variants in *PSAP* among *GBA* carriers may suggest oligogenic inheritance and that *PSAP* variants might act as genetic modifiers of risk in *GBA-*iRBD. This is in line with the biological link between sapC and GCase [6, 9]. In particular, it is possible that an impairment of the sapC-mediated activation of GCase contributes to an increased risk to develop iRBD in *GBA* variant carriers. These hypotheses require additional genetic and functional studies. We cannot rule out that the co-occurrence of *GBA* and *PSAP* variants is a coincidence, due to chance alone. However, the fact that two out of three extremely rare *PSAP* LoF variant carriers also carried a *GBA* mutation makes a coincidental association less likely.

It is still unclear whether *PSAP* mutations alone can increase the risk of iRBD or PD. It is possible that LoF of sapC, as seen in our patient with the p.332_333del mutation, will result in reduced activation of GCase and be an independent risk factor. On the other hand, it is also possible that *PSAP* variants might lead to iRBD through mechanisms independent of *GBA*. A possible mechanism can be due to an impairment of CTSD and progranulin (PGRN) activity, as previously hypothesized in PD [8]. PSAP, CTSD and PGRN interact in a network involved in lysosomal homeostasis and clearance of alpha-synuclein. PSAP dysfunction might lead to decreased transport of PGRN into the lysosome, reduction of the pro-CTSD conversion into active CTSD, and consequently to impaired lysosomal trafficking and degradation of deleterious or overrepresented proteins, such as alpha-synuclein [8].

This study has several limitations. Age and sex differed between patients and controls. However, this difference would generally lead to false negative results (as young mutation carriers still would not develop the disease), and is, therefore, less likely to affect our results, as no carriers were found in the controls. Although this study was performed in the largest genetic cohort of iRBD patients worldwide, the sample size may still be insufficient to detect extremely rare variants in *PSAP*. Finally, we were able to find *PSAP* LoF variants, different from each other, in only 3 iRBD patients. However, the absence of such variants in the ∼twofold larger control group and in the ∼140-fold larger gnomAD control population suggests that this finding might not be random.

Further studies in larger cohorts and functional analyses will be required to clarify the role of *PSAP* variants in iRBD and alpha-synuclein physiopathology. In addition, studies in other populations, such as East Asians, where *PSAP* variants have already been proposed as PD risk factors [10-14], will be necessary to further explore differences in the genetic underpinnings of synucleinopathies between different ethnic groups.

## Data Availability

All data is available upon reasonable request from the corresponding author.

## Acknowledgments

We wholeheartedly thank the participants in this study. This work was financially supported by the Michael J. Fox Foundation, Parkinson’s Society Canada, the Canadian Consortium on Neurodegeneration in Aging (CCNA), and the Canada First Research Excellence Fund (CFREF), awarded to McGill University for the Healthy Brains for Healthy Lives (HBHL) program. JFG holds a Canada Research Chair in Cognitive Decline in Pathological Aging. GAR holds a Canada Research Chair in Genetics of the Nervous System and the Wilder Penfield Chair in Neurosciences. WO is Hertie Senior Research Professor, supported by the Charitable Hertie Foundation, Frankfurt/Main, Germany. The Oxford Discovery study was funded by the Monument Trust Discovery Award from Parkinson’s UK and supported by the National Institute for Health Research (NIHR) Oxford Biomedical Research Centre based at Oxford University Hospitals NHS Trust and University of Oxford, the NIHR Clinical Research Network and the Dementias and Neurodegenerative Diseases Research Network (DeNDRoN). ZGO is supported by the Fonds de recherche du Québec - Santé (FRQS) Chercheurs-boursiers award, and is a William Dawson Scholar. We thank Daniel Rochefort, Helene Catoire and Vessela Zaharieva for their assistance.

